# Classification of Schizophrenic Traits in Transcriptions of Audio Spectra from Patient Literature: Artificial Intelligence Models Enhanced by Geometric Properties

**DOI:** 10.1101/2024.04.05.24305390

**Authors:** Paulo César F. Marques, Lucas Rafael F. Soares, André Victor de A. Araujo, Arthur Ribeiro Monteiro, Arthur Almeida Leitão Batista, Túlio Farias Pimentel, Lis de Lima Calheiros, Maria Helena N. S. Padilla, André Pacheco, Fabio Queda, João Ricardo M. Oliveira, José Luiz de Lima Filho, Silvana Bocanegra, Jones Albuquerque

**Affiliations:** Instituto Keizo Asami - iLIKA - UFPE; Centro de Ciências Médicas - UFPE; Faculdade de Medicina de Olinda - FMO; Departamento de Estatística e Informática - DEINFO - UFRPE; Departamento de Matemática - DMAT - UFPE; Centro de Informática - UFPE

**Keywords:** schizophrenia, classification models, classification models geometrics

## Abstract

Schizophrenia is a severe mental illness that affects approximately 1% of the global population and presents significant challenges for patients, families, and healthcare professionals. Characterized by symptoms such as delusions, hallucinations, disorganized speech or behavior, and cognitive impairment, this condition has an early onset and chronic trajectory, making it a debilitating challenge. Schizophrenia also imposes a substantial burden on society, exacerbated by the stigma associated with mental disorders. Technological advancements, such as computerized semantic, linguistic, and acoustic analyses, are revolutionizing the understanding and assessment of communication alterations, a significant aspect in various severe mental illnesses. Early and accurate diagnosis is crucial for improving prognosis and implementing appropriate treatments. In this context, the advancement of Artificial Intelligence (AI) has provided new perspectives for the treatment of schizophrenia, with machine learning techniques and natural language processing allowing a more detailed analysis of clinical, neurological, and behavioral data sets. The present article aims to present a proposal for computational models for the identification of schizophrenic traits in texts. The database used in this article was created with 139 excerpts of patients’ speeches reported in the book “Memories of My Nervous Disease” by German judge Daniel Paul Schreber, classifying them into three categories: 1 - schizophrenic, 2 - with schizophrenic traits and 3 - without any relation to the disorder. Of these speeches, 104 were used for training the models and the others 35 for validation.Three classification models were implemented using features based on geometric properties of graphs (number of vertices, number of cycles, girth, vertex of maximum degree, maximum clique size) and text entropy. Promising results were observed in the classification, with the Decision Tree-based model [1] achieving 100% accuracy, the KNN-k-Nearest Neighbor model observed with 62.8% accuracy, and the ‘centrality-based’ model with 59% precision. The high precision rates, observed when geometric properties are incorporated into Artificial Intelligence Models, suggest that the models can be improved to the point of capturing the language deviation traits that are indicative of schizophrenic disorders. In summary, this study paves the way for significant advances in the use of geometric properties in the field of psychiatry, offering a new data-based approach to the understanding and therapy of schizophrenia.

## Introduction

Schizophrenia is a severe mental illness that affects approximately 1% of the global population^1^, impacting an estimated 22 million lives worldwide^2^. It is a mental disorder characterized by a variety of symptoms, such as delusions, hallucinations, disorganized speech or behaviour, and cognitive impairment^3^. The early onset of this condition, coupled with its chronic trajectory, renders it a debilitating challenge for many patients and their families^2^. Disability often results from both negative symptoms (characterized by losses or deficits) and cognitive symptoms, such as impairments in attention, working memory, or executive function^4^. Furthermore, relapses can occur due to positive symptoms, such as paranoia, delusions, and hallucinations^2^. The inherent complexity of schizophrenia has led to a lack of consensus regarding the diagnostic, etiological, and pathophysiological criteria of the disorder^5^. The stigma associated with mental disorders often perpetuates additional challenges, such as difficulties in accessing employment, housing, and adequate healthcare^6^. An enhanced understanding of schizophrenia not only benefits individual treatment but also contributes to society in social and economic aspects, at the very least^7^.

Communication disorders are evident symptoms in various severe mental illnesses, presenting as a striking feature^8^. However, recent technological advancements have aided psychiatrists in understanding and assessing these changes in communication. Automated computerized semantic, linguistic, and acoustic analyses detail the ability to objectify and quantify communication patterns in a less subjective and biased manner present in any doctor-patient relations^9^. In diseases such as schizophrenia, bipolar disorder, and severe depression, both verbal and body expressions are affected. This can manifest in peculiar language patterns, changes in vocal intonation, and even unusual pauses during speech. The subjective interpretation of these patterns can lead to diagnosis variations^9^.

The differential diagnosis of psychosis is based exclusively on oral interview assessments with patients, an objective quantification of the speech disorders that characterize mania and schizophrenia. Efforts have been made to quantify the disorders through speech analysis^10^. A study conducted at the Danish Psychiatric Central Research Register with 300 cases concluded that 89.7% of the schizophrenia diagnoses were valid^11^. Another study conducted at the Finnish Hospital Discharge Register shows that schizophrenia diagnoses were validated with 74% for (DSM-IV) and 78% for (ICD-10)^12^.

In this context, technology has provided new perspectives for treating schizophrenia. Artificial intelligence techniques such as machine learning, natural language processing, and data analysis have allowed for more standardized and repeatable analysis of clinical, neurological, and behavioural data sets, with the potential to identify patterns that are sometimes not perceptible to psychiatrists^13^. The current diagnosis of schizophrenia is strongly linked to the physician’s experience^9^.

Machine learning algorithms, components of Artificial Intelligence, allow machines to learn through trial and error, functioning as a form of “alchemy”^14^. These algorithms can be seen as an “alien technology,” since there are no strict criteria for preferring one AI architecture over another, nor is it known why some algorithms work while others do not^14^. Some of the reasons for the failure of AIs are the poor quality of the data used for tool development, the distortion of the system’s absolute accuracy, and confusion in the origin of certain data sets, which distorts the training of models^15^.

The COVID-19 pandemic served as a significant test for AI and medicine, a time when many predictive tools were developed, but none were relevant, and some were potentially harmful to society^15^. In text analysis, despite the euphoria with the supposed success story of ChatGPT with medicine, AI is flawed because it is restricted to strictly syntactic aspects. This AI algorithm is not a competent and reliable source of information for clinical practice, as it cannot provide medical information according to the criteria of evidence-based medicine^16^.

Fortunately, Artificial Intelligence linked to geometry geometry can provide more accurate, safe, and reliable results. In recent years, Copelli et al.^17^ reported the possibility of correctly classifying negative symptoms and successfully diagnosing schizophrenia 6 months in advance through computational theoretical analysis of verbal report graphs^17,18^. This analysis of speech connectivity is capable of providing behavioural markers of formal thought disorders in psychosis (e.g., the disorganized flow of ideas), being of extreme importance as a complementary psychiatric assessment in chronic psychotic patients^17,18^. Classification based on automated analysis of semantic and syntactic speech combined with machine learning outperformed that based on clinical classifications, indicating that automated speech analysis can increase the predictive power of diagnosis beyond expert clinical opinion and predict the subsequent onset of psychosis in young people at high clinical risk^19^.

Unlike AI’s performance during the COVID-19 pandemic, the use of geometry allowed for the detection of early warning signs of the emergence of a possible new pandemic^20^. While AI naturally tends to accumulate machine learning error rates, geometric strategies, which use stable parameters, seem to capture better the “predictability” of phenomena with low linearity^21^. Therefore, the use of geometry linked to AI can anticipate the diagnosis of schizophrenia and the outbreak of crises in schizophrenic patients.

This work adds to the classical IA techniques’ geometric properties for classifying schizophrenic traits found in audio files of patients extracted from the literature, which was used as a Bench Test for the proposal presented here.

## Methods

Daniel Paul Schreber’s autobiography was chosen as a training base due to its relevance in clinical aspects of the disease schizophrenia, representing in the first person the discourse, delusions, hallucinations, experiences, clinical evolution, and sociocultural context of a patient with “dementia praecox,” now called schizophrenia.

The work differs from other autobiographies by reporting the experience of a schizophrenic patient still in the condition of the disease, while other works such as “The Center Cannot Hold: My Journey Through Madness” by Elyn R. Saks^22^ and “An Unquiet Mind” by author Kay Redfield Jamison^23^, bring the “sane” perspective after the occurrence of the patient’s outbreak moments. In addition, it is possible to identify the richness of details and testimonies lived by Schreber since he had a high degree of education, bringing in his work moments that transit between lucidity and reasonableness and delusions and hallucinations.

Schreber was a patient analyzed by Freud without a single face-to-face encounter, using only the words and experiences he brought in his book. Thus, the work contributed to the development of psychoanalysis since Freud referred to it in one of his literary works, addressing the etiological mechanism of paranoias through Schreber’s delusional discourse. Freud refers to “Memoirs of a Nervous Illness” as fundamental to the development of his first theory of psychosis.

In addition, the work transcends psychiatric reporting, advancing to a historical reference landmark. Constructing philosophical perspectives that led to recognising the Germans as a people and nation was essential.

### Database

The database used in this article was taken from the book *“Memoirs of a Nervous Illness”*, written by the German judge Daniel Paul Schreber. A total of 139 excerpts from this book were used, and these excerpts were divided and classified into three classes: 1 - excerpts with clear evidence of schizophrenia; 2 - excerpts with schizophrenic traits; and 3 - excerpts without any evidence related to schizophrenia, used here in this work as control cases. The 139 excerpts were labeled, with 51 (fifty-one) excerpts classified as schizophrenic (class 1); 47 (forty-seven) excerpts as having schizophrenic traits (class 2); and 41 (forty-one) excerpts as “Normal” or control cases (class 3). This database was divided using 104 as the training set and another 35 for validation and accuracy verification of the implemented models.

These texts were processed using scripts written in the Python programming language with the goal of extracting the characteristics that would be used in training the classification models. These characteristics are geometric properties contained in graphs (explained in the next section), the properties are: number of vertices, number of cycles, girth, vertex of maximum degree, maximum clique size, in addition to these properties, the text’s entropy was also calculated and used in the classification. The classifiers were implemented and trained using these properties of the graphs generated from the texts.

### Graph Theory

Graph theory is an area of mathematics that studies collections of objects and their relationships. A graph is a structure composed of a set of objects (vertices or nodes) that may or may not be connected (by edges or lines). This area has found various applications in medicine, providing powerful tools for analyzing and modelling a variety of clinical and biological problems. More precisely, a graph *G* = (*V, E*) is defined as a pair consisting of a finite set *V* and a set *E* ⊂ [*V*]^2^ of edges, where [*V*]^2^ denote the 2-element subsets of *V*. To avoid ambiguity, assuming that *V* ∩ *E* = ∅. The elements of *V* are called vertices, and the elements of *E* are the edges. By convention, we write *e* = *uv* for the edge *e* = {*u, v*} ∈ [*V*]^2^. The most common way to represent a graph is to draw a point for each vertex and connect two of these points with a line if they form an edge.

A subgraph *H* of a graph *G* = (*V, E*) is a graph formed by subsets of the vertices and edges of *G*, preserving the membership relations of *G*. Thus, 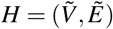, where 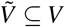 and 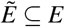, such that if 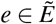, with *e* = *uv*, then *u*, 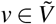. A path *C* linking the vertices *u* and *v* is a subgraph whose vertices form a sequence *v*_1_*v*_2_…*v*_*n*_ starting at *v*_1_ = *u* and ending *v*_*n*_ = *v*, and the edges are given by *e*_*i*_ = *v*_*i*_*v*_*i* −1_, for *i* = 1, 2, …, *n* − 1. When, for any 1 ≤ *i < j* ≤ *n*, we have that *v*_*i*_ ≠ *v*_*j*_, we say that the path is simple. Furthermore, if *v*_1_ = *v*_*n*_, we have a cycle^24^.

### Graph from Text

Given a text composed of words, we transform it into a graph, such that each word *P* is modelled as a vertex of the graph and each word *A* preceding *P* or *C* following *P* in the text becomes an edge (*P, A*) or (*P,C*) in the graph (does not generate parallel edges). For example, if we have a sequence of words *R, P*, and *Q*, the graph will be formed by vertices *R, P*, and *Q* and edges (*R, P*) and (*P, Q*). [10]. It is worth noting that this algorithm does not repeat words for the vertices, nor does it include special characters, commas, periods, and does not differentiate between lowercase or uppercase. In this way, each word will appear only once in the graph, and every time a word in the text is repeated (not neighbouring), a cycle is formed.

If all n words are distinct, then we have a simple path with n1 edges. If all n words are the same, then we have a vertex and a loop (an edge starting and ending at the same vertex).

When the graph is generated, as described previously, some information about it is generated, such as the number of vertices, which is the number of non-repeated words, and the degree of each vertex. With this, we know how many neighbours each word has, the number of cycles, the size of the smallest cycle (girth), and some other information that can be extracted from the graph.

In addition to the exposed properties, graphs allow us to calculate other characteristics that can be used to characterize the graph generated from that text. Centrality is one of these characteristics. Centrality, in a graph, is a measure that establishes how important a vertex is in the graph. In theory, the most known measures for centrality are degree centrality, betweenness centrality, closeness centrality, and eigenvector centrality^25^. For example, betweenness centrality quantifies the frequency with which a node acts as a bridge along the shortest path between other nodes in the network. In other words, a node with high betweenness centrality acts as a vital hub for communication between different parts of the network. By definition, the betweenness centrality of a node v in a network G is calculated as the sum of the fraction of all pairs of nodes s and t that have v on the shortest path between them. This is formally expressed as

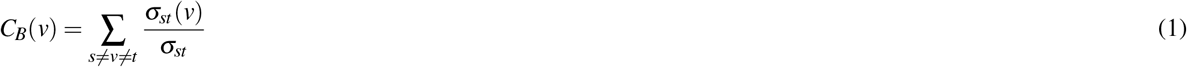

- *C*_*B*_(*v*) is the betweenness centrality of node *v*.
- *σ*_*st*_ is the total number of shortest paths from *s* to *t*.
- *σ*_*st*_(*v*) is the number of shortest paths from *s* to *t* that pass through node *v*.

Based on the centralities (equation(1)), as mentioned above, obtained from the graphs generated from the texts of the 3 classes, a model (referred to later as model [3]) based on centrality metrics for text classification was implemented.

### Information Theory

Information theory is applicable because it is based solely on the probability distribution associated with one or more variables. By using the probability distributions associated with the values of the variables to check whether they are correlated or not, and depending on the situation and how the correlation is established, it can be applied in linear or non-linear systems.

Information theory quantities involving one and two variables are well-defined and their results are well-understood^26^. With respect to the probability distribution *p*(*x*) of a variable *X*, the canonical measure is the Shannon entropy *H*(*x*) given by:

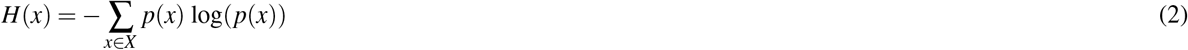

and measures the amount of uncertainty present in the probability distribution.

As it is defined, when the probability distribution is concentrated near one value, the entropy will be low, while in the case of a uniform probability distribution, the entropy will be maximized.

### Decision Tree Classification

A decision tree is a non-parametric supervised machine learning method used for classification and regression with the goal of creating a model that predicts the value of a target variable by learning simple decision rules inferred from the data features. A tree can be seen as a piecewise constant approximation.

Thus, based on the characteristics extracted from the graphs (mentioned in the database section) generated from the texts of the 3 classes, a model (referred to hereafter as model [1]) based on Decision Tree for text classification was implemented.

### K - Nearest Neighbors

K-Nearest Neighbours is a supervised machine learning method based on neighbours. It is divided into two types: classification for data with discrete labels and regression for data with continuous labels. The method for discrete labels was used for the implemented model. The principle behind the nearest neighbour method is to find a predefined number of training samples closest to the new point and predict the label from them. The number of samples can be a constant defined by the user (k-nearest neighbour learning).

To calculate the best K to be used, the best score was calculated for each K varying from 1 to 30. The number 5 was found to be the best for K as shown in Figure 4.

**Figure 1.**
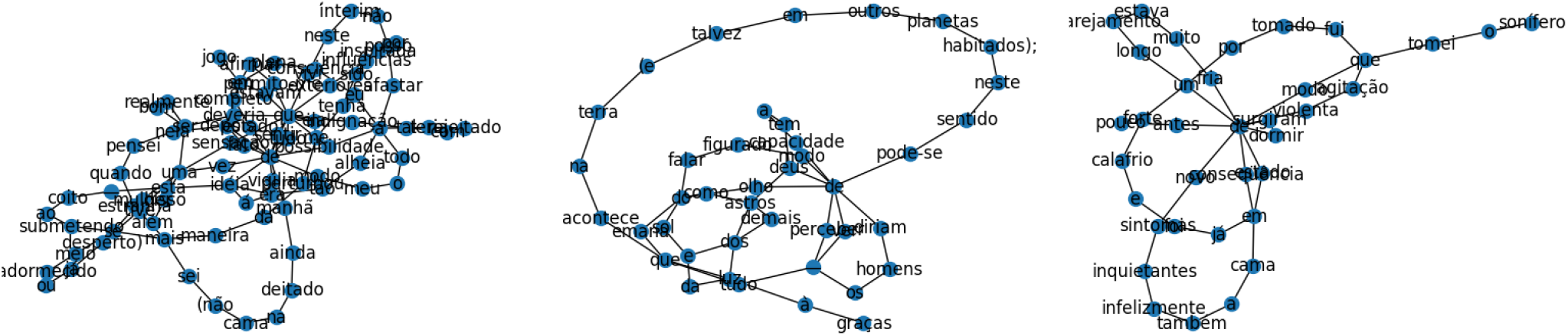
Graphs generated from texts a: Schizophrenic text; b: text that presents Schizophrenic traits; and c: control group text. Source: elaborated by the authors.

**Figure 2.**
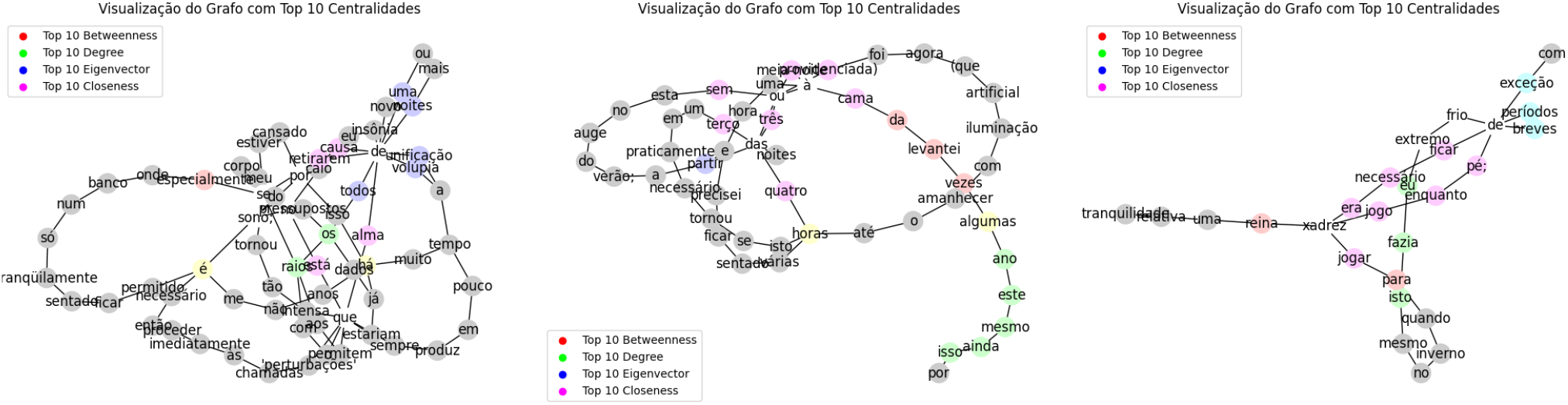
Calculation of centralities in graphs a: Schizophrenic text; b: text that presents Schizophrenic traits; and c: control group text. Source: elaborated by the authors.

**Figure 3.**
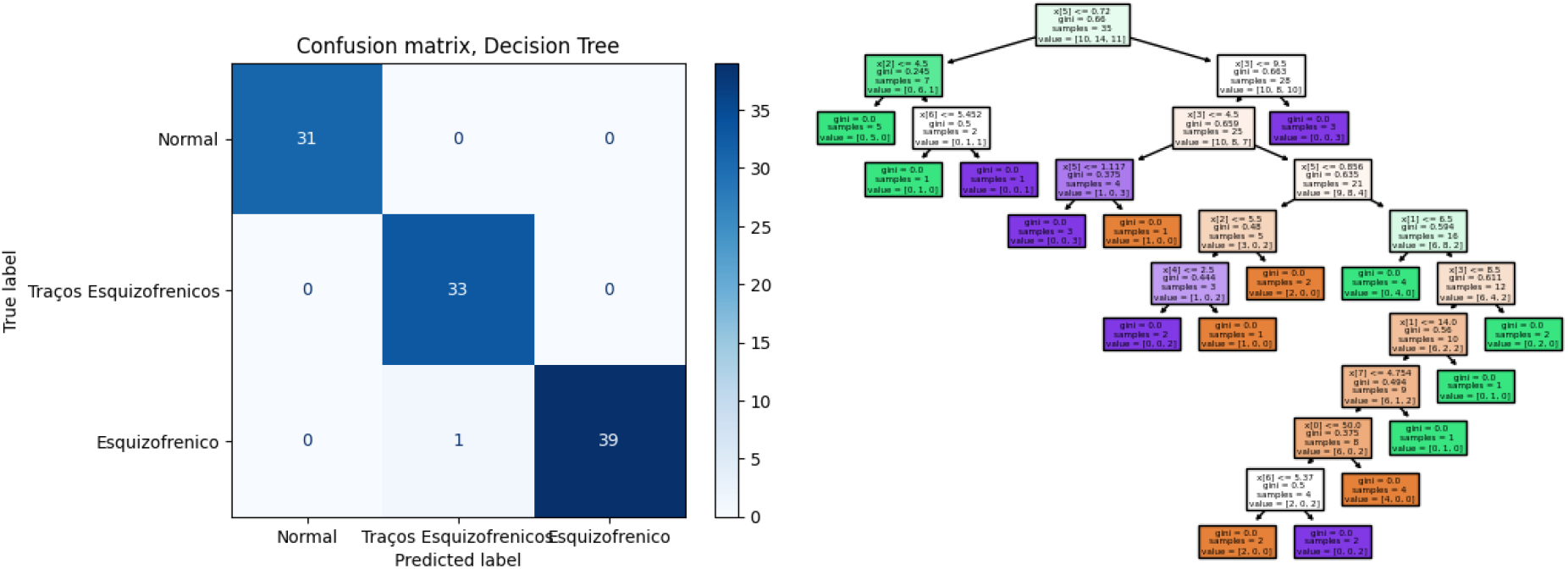
Implemented decision tree confusion matrix; Representation of the decision tree trained with texts. Source: elaborated by the authors.

**Figure 4.**
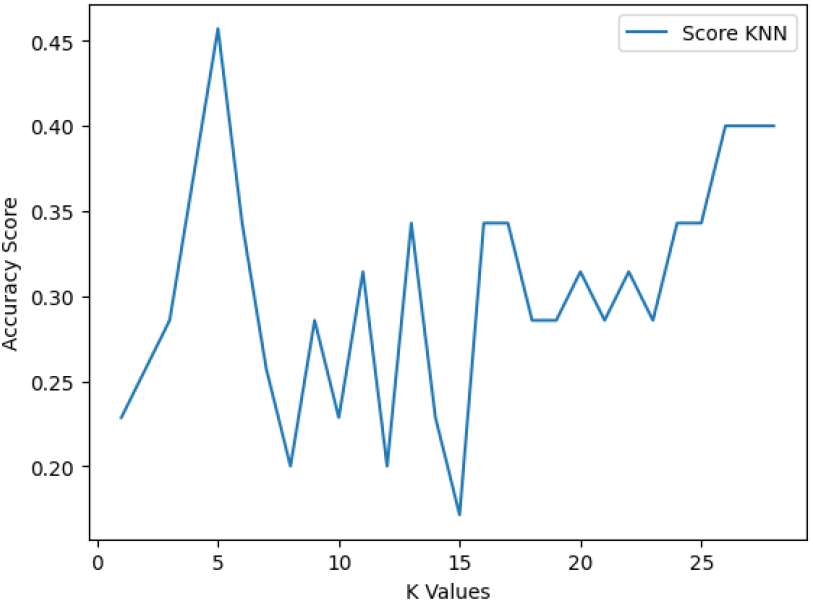
K-NN Score. Source: elaborated by the authors.

Thus, based on the characteristics extracted from the graphs (mentioned in the database section) in addition to the entropy of the text from the 3 classes, a model (referred to as model [2] in the text) based on K-NN (5-NN) for text classification was also implemented.

## Results

The results were divided into model [1], model [2], and model [3]. The performance metrics selected to evaluate the effectiveness of the models were [a] accuracy, [b] precision, and [c] recall.

After training models [1] and [2] using 104 texts where 31 (thirty-one) excerpts were labelled as “Normal” or control cases, 33 (thirty-three) excerpts presented schizophrenic traits and 40 (forty) excerpts were classified as schizophrenic, another 35 (thirty-five) texts were classified to obtain validation of the models.

The metrics for model [1] reached a precision [b] of 100%, a recall [c] of 100%, and an accuracy [a] of 100% as shown in Figure 5. These results suggest that the model has a very high rate of correct prediction (precision) while maintaining a substantial accuracy of true positive results among all relevant positive predictions [c].

**Figure 5.**
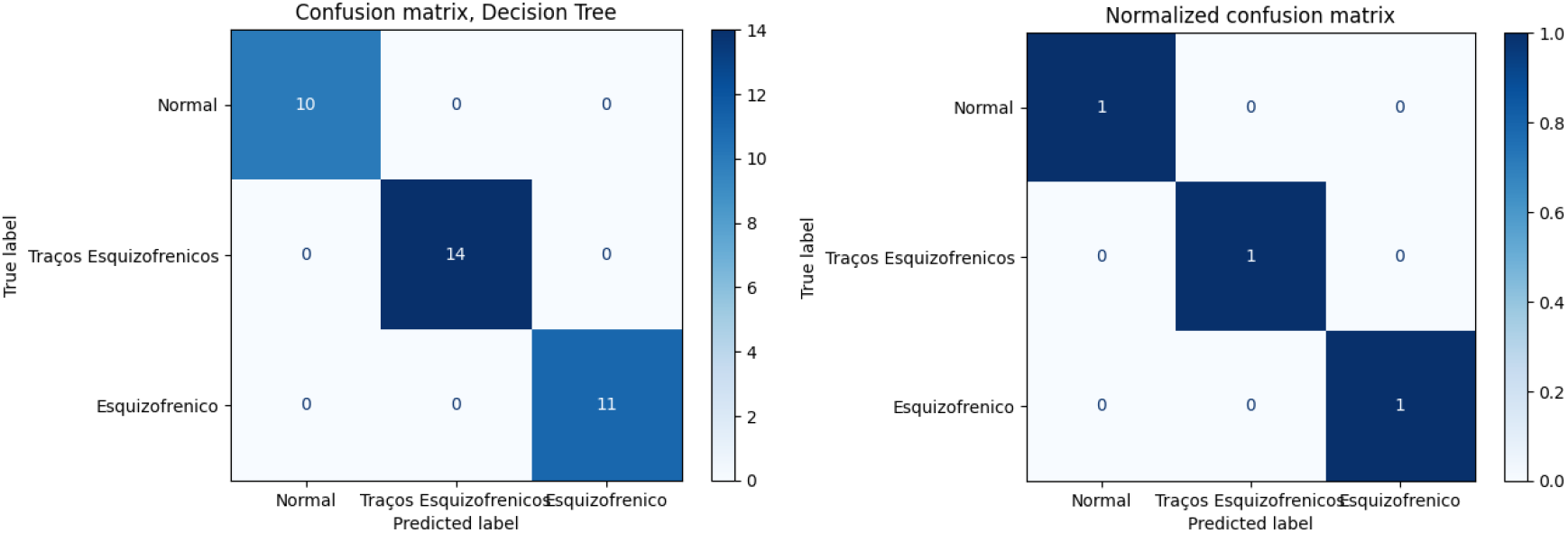
Decision Tree Confusion Matrix. Source: elaborated by the authors

For model [2], a precision [b] of 66%, a recall [c] of 62.8%, and an accuracy [a] of 62.8% were achieved, as seen in the confusion matrix in Figure 6.

**Figure 6.**
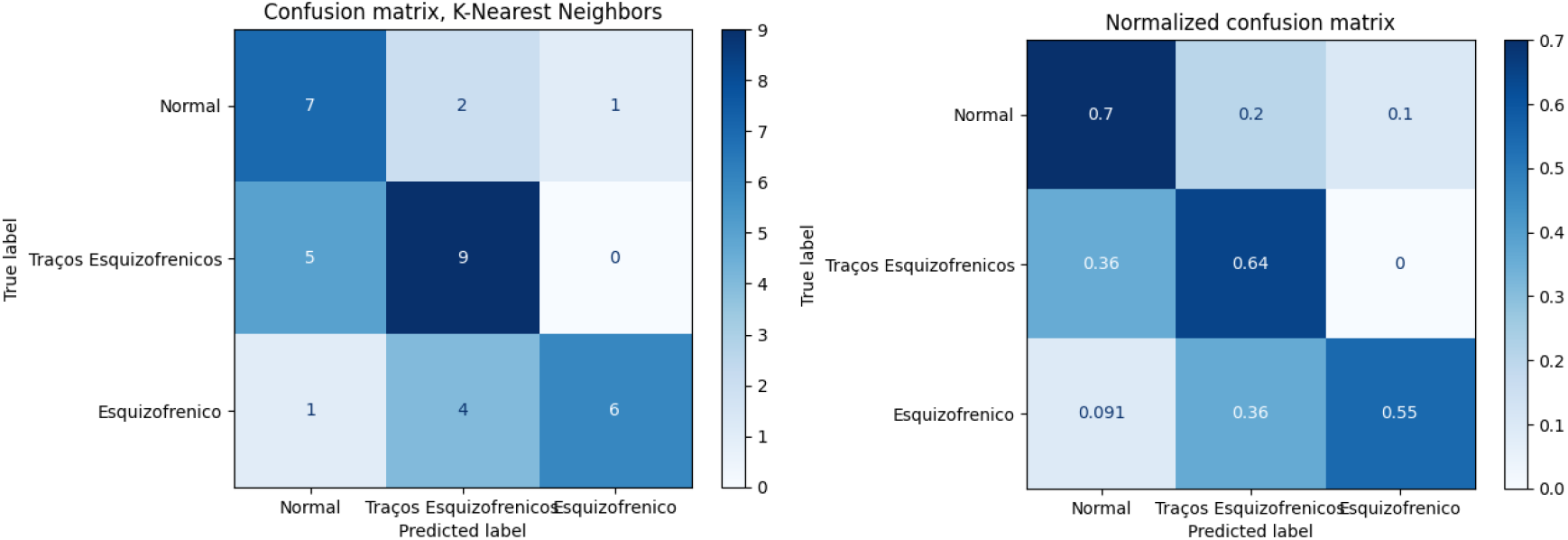
Confusion Matrix for KNN. Source: elaborated by the authors

**Figure 7.**
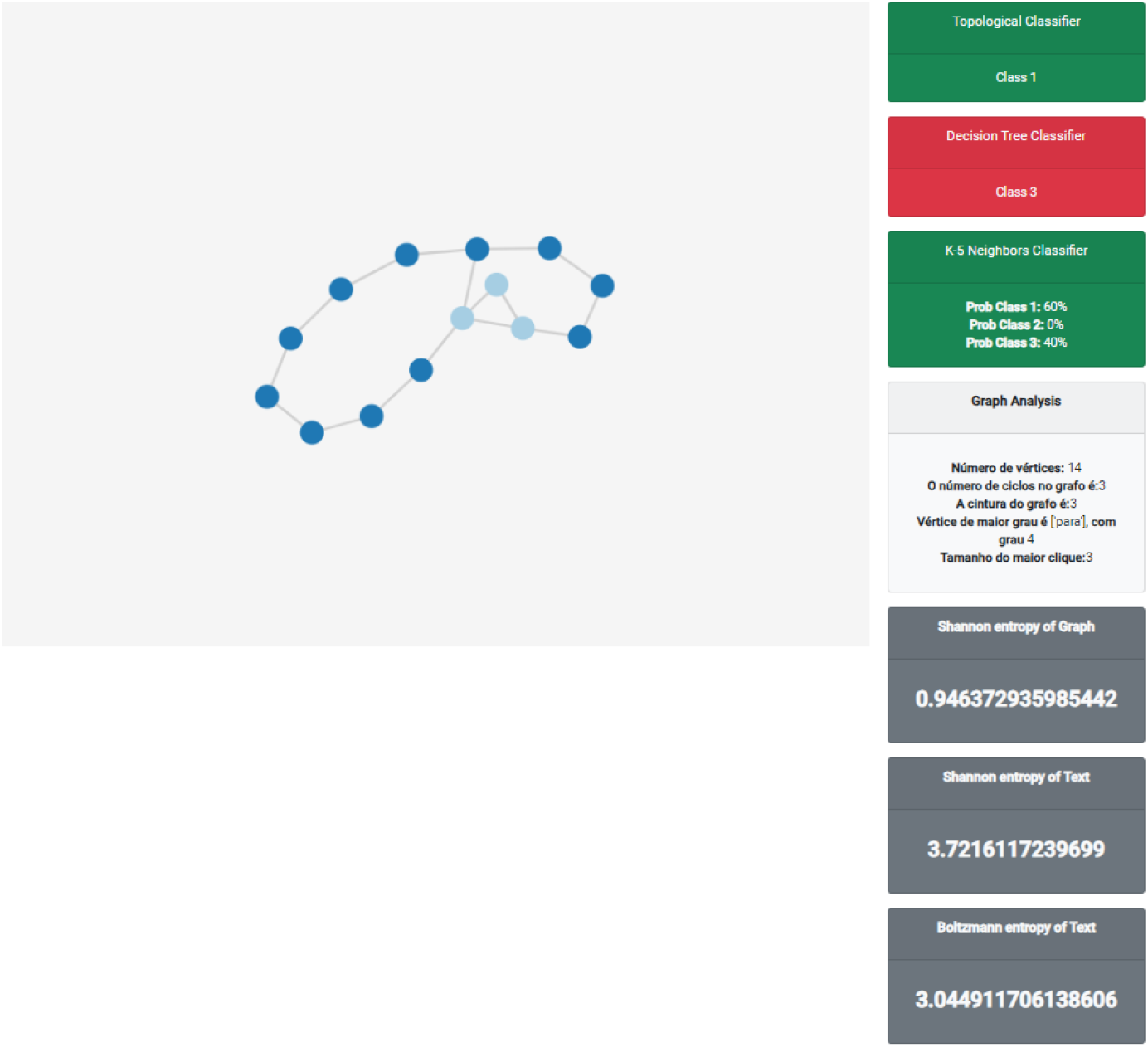
Practical implementation. Source: elaborated by the authors

A qualitative analysis of the classifications reveals that model [1] is exceptionally competent in identifying and classifying texts using geometric properties of graphs, and information entropy. However, the limitations of the model [2] became evident when labeling the texts. Therefore, another model based solely on graph centralities, model [3], was implemented. However, the precision was below expectations, achieving a precision [b] of 59% for schizophrenic texts and a precision [b] of 63% for the control group.

In summary, the results are promising and indicate that using graph properties and entropy to classify schizophrenic text offers a viable path for practical applications. However, we recognize the need for continuous improvement, particularly regarding handling ambiguous texts and balancing datasets.

In addition to the classification, an application for real-time text classification was implemented. It can be accessed from a browser at the following address: https://www.healthdrones.science/schizophrenia/SpeechGraph/

## Discussion

The [1] decision tree-based classification model demonstrated remarkable performance, achieving 100% in all performance metrics: precision, recall, and accuracy. This indicates that the model is highly efficient in correctly predicting classifications, maintaining a high rate of true positives among all relevant optimistic predictions. However, it’s essential to consider that a model with 100% in all metrics may indicate potential overfitting. Overfitting occurs when the model fits the training data so well that it cannot generalize adequately to new data. Therefore, conducting additional tests with new data would be beneficial to confirm the model’s efficacy.

On the other hand, the model [2] K-NN model showed limitations, achieving a precision of 66%, a recall of 62.8%, and an accuracy of 62.8%. This could be attributed to the nature of the K-NN algorithm, which is sensitive to the choice of the number of neighbours (K) and the distance metric used. Furthermore, K-NN may struggle with high-dimensional data due to the “curse of dimensionality.” Thus, it would be interesting to explore different configurations of the K-NN algorithm to improve its performance. Additionally, a third model [3] based solely on graph centralities exhibited lower-than-expected precision, with 59% for schizophrenic texts and 63% for the control group. This suggests that graph centrality alone may not be sufficient for effective text classification. Combining graph centralities with other features may enhance the model’s performance.

Although the results are promising, they also highlight the need for ongoing research and development to overcome the limitations of existing models and improve classification accuracy. This includes exploring different algorithm configurations, combining various features, and conducting more tests with new data. Moreover, it is crucial to consider issues such as handling ambiguous texts and balancing datasets.

Our findings in this work have some implications. The assertive identification of patients at risk of developing schizophrenia through speech analysis by increasing predictive power would be extremely useful in early psychiatric intervention and improving prognosis. Furthermore, the computerized analysis of complex human behaviours, such as speech, can elevate the level of psychiatry, allowing for a more objective and reliable clinical analysis. With the automated evaluation of speech, which will function as a kind of “laboratory test” and complement the clinical examination, doctors will have access to previously inaccessible information, enabling better treatment decisions and prognosis.

## Conclusion

In this study, we explored the implementation of text classification models for identifying linguistic patterns associated with schizophrenia. The results demonstrate these models’ feasibility and effectiveness in distinguishing texts with schizophrenic traits from normal texts„ achieving a precision of 100% in one of the models (model [1]). This high precision rate suggests that the models can capture subtle nuances in language indicative of schizophrenic disorders.

The application of these models has the potential to revolutionize the way mental health professionals approach the diagnosis and monitoring of schizophrenia. By providing an objective and quantitative tool for assessing patients’ language, the models can assist in the early detection of schizophrenic symptoms, as well as in monitoring the progression of the disease and the response to treatment.

For future work, we plan to expand our research to clinical settings, conducting interviews with schizophrenic patients and applying the implemented models to analyze their language patterns in real time. This will allow us to further validate the effectiveness of the models in a practical context and explore their potential for assisting in the diagnosis and personalized treatment of schizophrenia. In summary, this study paves the way for significant advancements in the field of psychiatry, offering a new data-driven approach to understanding and managing schizophrenia. With the continued evolution and validation of these text classification models, we are one step closer to an era where the diagnosis and treatment of schizophrenia are more accurate, efficient, and personalized.

## Data Availability

All data produced in the present study are available upon reasonable request to the authors

## Acknowledgements

This work is partially supported by CNPq-IA2 grant 409818/2022-4.

## Notes

### Competing Interest Statement

The authors have declared no competing interest.

